# Mapping rehabilitation pathways after cardiac surgery: Identifying key points for patient involvement and gaps in care

**DOI:** 10.1101/2025.04.29.25326494

**Authors:** Bente S Toft, Hilary L Bekker, Lotte Ø Rodkjær, Ivy S Modrau

**Affiliations:** Department of Clinical Medicine, Cardiothoracic Surgery, Aarhus University, Aarhus, Denmark; Department of Cardiothoracic and Vascular Surgery, Aarhus University Hospital, Aarhus, Denmark; School of Medicine, University of Leeds, Leeds, UK; Leeds Unit of Complex Intervention Development (@LUCID_Leeds), Leeds, UK; Department of Public Health, Aarhus University, Aarhus, Denmark; Department of Infectious Diseases, Aarhus University Hospital, Aarhus, Denmark

**Author notes:** Corresponding author: **E-mail:** **(BST)**.

**Keywords:** Qualitative research, health services research, document analysis, cardiac rehabilitation, cardiac surgical procedures, patient participation, Denmark

## Abstract

**Background:** Cardiac rehabilitation in Denmark is cross-sectional and decentralised to municipalities, resulting in different pathways and practices. A comprehensive overview of these variations and their impact on the rehabilitation process following cardiac surgery is lacking.

**Aim:** This study aims to map the rehabilitation pathways after cardiac surgery, and to identify key interactions between patients and clinicians that influence rehabilitation outcomes and patient experiences.

**Methods:** A qualitative multi-method study was conducted using metro mapping methods, including document analysis, stakeholder consultations, practice observations and go-along interviews across multiple sites in Central and Northern Denmark Regions.

**Results:** Mapping revealed significant variability in cardiac rehabilitation pathways across healthcare settings, including diverse referral, enrolment, content, delivery methods, and follow-up procedures, involving multiple clinicians. A consistent finding across all pathways was both a prolonged transition from in-hospital phase 1 to outpatient phase 2 rehabilitation, resulting in a gap of care, and minimal patient involvement in decision-making and planning during the early phase of recovery.

**Conclusion:** This study provides a comprehensive overview of current rehabilitation pathways following cardiac surgery, revealing significant variability and challenges. Our findings emphasise the need for a person-centred redesign of the cardiac rehabilitation pathway with improved sector transitions. Our findings provide guidance for clinicians and underscore the importance of clear legal frameworks, consistent guidelines, and co-design of cardiac rehabilitation programmes with patients. Strengthening these areas could promote greater patient engagement, reduce variability in service delivery, and ultimately lead to better patient outcomes and experiences.

## Introduction

Cardiac rehabilitation (CR) is highly recommended and an integrated part of the patient pathway after cardiac surgery. CR improves postoperative outcomes including reduced mortality, fewer hospital readmissions, and improved quality of life after coronary artery bypass grafting and heart valve surgery (1).

In Denmark, all patients are entitled to rehabilitation after cardiac surgery, fully funded by the tax-financed public healthcare system (2). According to the Danish CR database, 61% of all cardiac patients participate in the physical training component of CR after discharge (3). No data are available on participation rates specifically among cardiac surgery patients, who may face greater barriers to engagement due to the complexity of their recovery, indicating potential for improvement. Moreover, timely intervention and seamless coordination across sectors are critical to maintain participation and adherence to the CR programmes (4–8).

CR pathways consist of three phases: phase 1 (in-hospital recovery), phase 2 (outpatient rehabilitation), and phase 3 (long-term maintenance) ensuring continuity of care and improvements along the pathway involving multiple sectors and a wide range of clinicians. In Denmark, the majority of patients receive Phase 2 CR as an outpatient service provided by municipalities. At this stage of the pathway, there are no national standards for clinicians, programme content, or duration, leading to likely variations in practice across municipalities. A key challenge is ensuring effective rehabilitation for patients with diverse recovery trajectories and varying physical, existential, and mental needs (9, 10). Currently, the guidelines of the European Society of Cardiology are the primary reference for CR in Denmark. It recommends individualized, collaborative CR programmes to improve participation, outcomes, and health equity (11). However, evidence on how current CR programmes incorporate patient and relative perspectives remains limited (9, 12–17).

This research study aims to map the rehabilitation pathways after cardiac surgery, and to identify key interactions between patients and clinicians that influence rehabilitation outcomes and patient experiences.

Meaningful patient involvement in healthcare innovation requires a clear understanding of patient pathways, the contextual environment, key points of interaction, and systemic barriers and it is crucial to identify which aspects of a complex intervention are effective, what needs improvement, and how changes can be feasibly implemented within the existing service context (18, 19).

This study will provide essential evidence of the context within which CR is delivered, to inform the development of interventions that enhance patient involvement in their healthcare and influence CR participation, effectiveness and accessibility (19).

## Methods

The study employs a qualitative exploratory design with a multi-method approach (20). It is part of a larger programme of work to develop an intervention aimed at advancing patient engagement with CR in Denmark, guided by the complex intervention framework for intervention development and evaluation (19). Metro mapping methods are used to visualize service delivery and the patient’s rehabilitation pathway, identifying key clinician-patient touchpoints (21). This information is used to promote a shared understanding of the CR pathway between patients and clinicians, across service providers.

Data collection involved inductive and iterative processes in two regions of Denmark, utilizing the following methods:

Document analysis focused on materials identified by key stakeholders as central to their practice and outlining the core components of their rehabilitation services. Both paper and electronic documents were included, and analysis continued until redundancy was reached (22).

Field observations and go-along interviews (23) were conducted in cardiothoracic hospital units, outpatient clinics, and municipal CR facilities for patients who had undergone cardiac surgery in two Danish regions. The municipalities were selected to represent diverse geographical locations, sizes, and populations (Table 1). A convenience sample of clinicians and the patients were observed in their clinical environment to identify contextual variations, actions, and interactions in activities relevant to CR. Ad hoc questions were asked about the pathway, their experiences and their perspectives on CR (24, 25).

**Table 1:** Consultations with key stakeholders from different settings.

| Setting | Nurses | Physio-therapists | Managers | Other professions |
| --- | --- | --- | --- | --- |
| Cardiac-thoracic surgery departments (n=2) of the Northern Denmark Region and Central Denmark Region | 6 | 3 | 2 | Social and health assistant (n=1)<br>Chaplain (n=1)<br>Surgeons (n=3)<br>Secretary (n=1)<br>Dietician (n=1)<br>Volunteers (n=3)<br>Sports instructor (n=1)<br>Politician (n=1)<br>General practitioner (n=1)<br>Researchers (n=8) |
| Outpatient clinics (n=3) | 4 | 3 | 1 |  |
| Regional hospital (n=1) | 1 | 1 | 0 |  |
| Municipalities (n=10)<br>urban (n=2)<br>intermediate (n=2)<br>rural (n=3)<br>remote (n=3). | 10 | 6 | 3 |  |
| Total | 21 | 13 | 6 | 21 |

Stakeholder consultations were conducted with a purposeful sample of multidisciplinary clinicians and CR experts (Table 1), who were contacted by the first author. The aim of these meetings, conducted in person, online, or by telephone, was to assess strengths, challenges, and needs, as well as to map CR pathways in specific settings.

Field notes were recorded in a diary format during and immediately after the observations and consultations, then transcribed as data (26).

All data collection was conducted by the first author, a postdoctoral researcher and trained physiotherapist with expertise in rehabilitation and qualitative methods. She had personal experience as the spouse of a cardiac surgery and rehabilitation patient but no prior relationship with any of the participants.

### Patient research partner involvement

This study involved five patient research partners to provide experienced-based knowledge and validate the data used to map CR pathways. The group was purposively selected to represent cardiac surgery patients with different age, gender, social background, disease type, recovery pathway, and municipality setting characteristics. The level and methods of patient research partner involvement, their contribution and their influence are described in a matrix (27)(S3 File).

### Ethical approval and consent to participate

The study was registered as an internal inventory of research projects with the Danish Data Protection Agency, Central Denmark Region, Regional Office (Skottenborg 26, DK-8800 Viborg), and the registration was confirmed (no. 1-16-02-304-24, 19 August 2024).

The Committees on Health Research Ethics of the Central Denmark Region (Regional Secretariat Legal Office, Skottenborg 26, DK-8800 Viborg) were contacted and confirmed that, in accordance with the Consolidation Act on Research Ethics Review of Health Research Projects, Consolidation Act No. 1268 of 28 November 2024, Section 14(1), the study could be conducted without approval from the Committees. Ethical oversight was therefore waived by the ethics committee.

Permission for observations and access to stakeholders was obtained from department managers. The study was conducted in accordance with the Helsinki Declaration (59). All participants were informed about the research project and volunteered to participate by giving verbal informed consent to be observed or talk to the researcher. Their anonymity and confidentiality were guaranteed. No financial incentives were provided, except for patient research partners, who were compensated for transport, remuneration, and catering.

### Data analysis

All data were interpreted by using the method of constant focused comparison, which aims to identify similarities and differences in context, actions and interactions related to rehabilitation (28). Metro map sketches were produced informed by each of the data collection methods for different pathways across the two regions in an iterative process of accumulating input.

Data were organised using the five layers of metro mapping: 1) experiences of key stakeholders and patient partners; 2) metro lines, colour-coded to visualize the timeline across the three rehabilitation phases, with symbols indicating key touchpoints where patients engage with clinicians; 3) information on activities during clinical encounters; 4) key clinicians involved, and 5) context of the encounter (21, 28, 29).

A prototype metro map was created using Microsoft Visio professional 2016© (Microsoft Cooperation, Redmond, WA, USA).

The study followed Standards of Reporting Qualitative Research (SRQR)(S1 File) and Guidance for Reporting Involvement of Patients and the Public in Research-Short Form (GRIPP2-SF) guidelines (S2 File)(30, 31).

## Results

Data collection was conducted from September 2023 to August 2024. A total of 72 documents were analysed and categorised as follows: reports and clinical guidelines (n=25), materials for patients and relatives (e.g., information letters, leaflets, and programme descriptions) (n=23), patient-reported outcome measures, dialogue tools, and screening instruments (n=17), and digital solutions (n=7). The documents addressed aspects and processes of care and provided guidance on CR practices.

Field observations and go-along interviews were conducted over 18 hours in seven different settings. Key activities observed included preoperative risk assessments and preparatory consultations, mobilisation sessions, discharge planning, telephone follow-ups, phase 2 group exercise programmes, ergometer bike testing, and patient education sessions.

Stakeholder consultations with 61 key individuals provided insights into the context and procedures of CR in different settings (Table 1).

### Metro mapping of CR pathway

The analysis of CR pathways revealed a complex and heterogeneous structure, that varied significantly between settings. Nurses and physiotherapists emerged as key clinicians delivering CR.

An overview of common CR pathways after surgery in two Danish regions is presented in metro maps (Fig 1). The pathways and touchpoints differed in both number and type of activities, depending on the discharging hospital, the subsequent municipalities involved, and in the individual patient’s risk profiles.

**Fig 1.** Metro maps of synthesised metro lines of Central and Northern Central Denmark regions including 5 layers (experience; context; activities, time and key providers).

### In-hospital phase 1 rehabilitation

In the absence of applicable national guidelines, the provision of information and exercise programmes regarding rehabilitation activities varied between the two cardiac surgery units. One hospital recommended general physical restrictions for the upper extremities for eight weeks, while the other allowed arm use within the “Keep Your Move in the Tube” principle, with restrictions lasting six weeks (32)

Short hospital stays, particularly due to early transfers to regional hospitals, limited the time available to build therapeutic relationships and understand patients’ life situations. Initial recovery on the cardiothoracic unit focused on physical care, including medication administration, wound care, mobility support, and vital sign monitoring. According to the clinicians neither hospital offered prehabilitation services for elective patients.

The emphasis on physical recovery often overshadowed other aspects of patient care:

“We are very much like a ‘body-factory’ and tell the patients, ‘And then you just do this,’ but this is no ‘just’ for the patients.” (Hospital nurse, NDR) When patients needed psychological and emotional support, nurses were the key providers, with occasional referrals to the Danish Heart Association or the hospital chaplain.

### Development and communication of CR plans

Nurses and physiotherapists provided limited information about CR, with patient consent typically obtained at discharge and few refusals. Standard care did not include systematic discussions with patients or their relatives about the rehabilitation pathway or preferences. Both hospitals provided discharge letters with rehabilitation information, but content and delivery varied. Verbal information often focused on physical recovery and restrictions, sometimes misaligned with written materials, leading to inconsistencies due to uncertainty in clinician communication. Doctors, responsible for the rehabilitation plan and referral, rarely interacted directly with patients regarding rehabilitation, mainly collaborating with physiotherapists on physical limitations or monitoring needs. Clinicians expressed uncertainty about what their colleagues were communicating to patients. Referral letters were considered more useful and complete regarding physical limitations and relevant psychosocial aspects when prepared by experienced clinicians. Delayed or missed referrals were attributed to irregularities in hospital processes, prolonged patient admissions, or sometimes unknown reasons. To improve the patient pathway across sectors, one region had appointed a designated CR coordinator. Phase 2 rehabilitation clinicians identified the lack of timely referrals and follow-up as key areas for improvement.

### Variations in service delivery procedures

CR referrals in the two regions studied were governed by different legal and financial frameworks. These frameworks resulted in varying referral procedures and management guidance. Under §140, municipalities are required to contact patients within seven days of receiving a referral plan, establish a clear timeline for starting phase 2 rehabilitation, and provide transport support. In contrast, §119 is more flexible and allows rehabilitation to be tailored to the patient’s needs in terms of timing, location, and type but does not guarantee the time of contact or transport assistance.

Under §86, individual inpatient rehabilitation in CR units was provided to patients with reduced functional capacity or special needs. Sometimes, clinicians criticized the §119 referrals, as they often needed to modify or combine the legal clauses:

“There are no advantages in using §119” (CR coordinator, Northern Denmark Region).

In one region, ‘high-risk patients’ were offered a specialised, intensively monitored, exercise-based group programme on an outpatient basis at the hospital, while in the other region, individual physiotherapy sessions were provided. The municipalities relied on different regional guidelines and pathway programmes, such as those targeting chronic diseases, addressing social inequalities, or establishing local cooperative agreements, and there was considerable variation in health service provision, including the content and duration of rehabilitation programmes.

### Care gap between phase 1 and phase 2

A significant care gap was identified between hospital discharge and phase 2 CR enrolment, characterised by a lack of structured support. During this period, patients were expected to manage recovery independently, with support from diverse materials such as mobile health (mHealth) applications, leaflets, and exercise programmes. Most patients had no formal follow-up until enrolling phase 2 out-patient rehabilitation programme. Relatives were perceived to receive little support during this time, despite the importance of their role:

“It is often worse for the relatives” (municipal nurse, NDR).

Handover processes were unclear, with no consistent responsibility for guiding patients through the rehabilitation pathway after discharge. Some patients received follow-up calls from a hospital nurse or were contacted by municipality clinicians and offered early follow-up, such as psycho-social and emotional support or physical activity guidance to prevent physical decline.

Outpatient phase 2 CR was often delayed six to eight weeks after surgery due to sternotomy restrictions, waiting lists, scheduling, or holiday closures. A notable disagreement among CR professionals regarding the optimal timing of initiation was observed. Some advocated for early support and exercises to enhance health and well-being, while others argued that, due to fatigue and physical limitations, most patients were not ready until six weeks post-surgery, raising concerns about the resource efficiency of early programmes. Patients not enrolled in standard group-based rehabilitation were at risk of missing psychosocial support, with follow-up typically limited to suture removal and routine consultations by general practitioners (GP). GPs were generally considered primarily responsible for coordinating care and providing counselling for stress, anxiety, or depression in these patients.

Delayed or missed referrals resulted in patients frequently contacting municipal rehabilitation services, where clinicians faced a high volume of questions and unmet needs. These concerns typically centred around wound healing, physical restrictions, activity guidelines, and psychosocial issues from both patients and their relatives. An unknown number of patients and their relatives may have needed additional support but did not seek help, leaving their needs unmet.

### Phase 2 rehabilitation

The transition to phase 2 CR and timely initiation of contact was seen as critical to enrolment: “’What do I need it for?’ asked the capable patients, when several weeks had passed before they were called” (hospital nurse, CDR).

In some cases, patients were unreachable or had reconsidered their decision to participate in rehabilitation and ultimately declined the offer.

Standard group-based CR programmes were well established in the municipalities. However, in rural areas, some municipalities did not have enough cardiac surgery patients to form groups, so alternative delivery formats or waiting time were required. Although the core elements of CR (exercise training, education on lifestyle changes, psychological support, risk factor management, and self-management) are well-defined, the programs varied significantly in design, structure, content, activity components, and duration. The procedures, methods, and materials used were not uniform.

A common feature of most programmes was an initial assessment typically involving a clarifying conversation (in-person or by telephone), often combined with patient-reported outcome measures. During this conversation, patients’ needs, health literacy, motivation, and vulnerability were assessed using a range of methods and tools, with screening for anxiety and depression using standard questionnaires. In some municipalities goal setting and CR planning were part of this conversation.

All but one of the local municipalities carried out repeated physical tests such as the six-minute walk-test combined with sit-to-stand-test or bike test at both the beginning and end of the course. The length of the courses and the time between tests varied. These conversations and tests enabled clinicians to assess patient perspectives and provided data for the Danish CR Database.

The flexibility of CR planning in clinical practice was shaped by local health policies and management priorities. In some instances, clinicians adapted the rules to better accommodate patient needs:

‘When we extend the courses, it goes under the radar’ (Municipality Nurse, NDR).

However, CR provision was often challenged by factors such as multimorbidity and changes in patients’ health or life circumstances. As part of a tailored approach, some patients were offered to participate in only selected elements of the standard programme. The most individualised courses included one-to-one sessions, home visits, or mHealth solutions for those unable or unwilling to take part in group-based activities.

The involvement and support of relatives during phase 2 CR varied widely, ranging from no involvement to special sessions or individual consultations for families and carers.

Supporting the transition to phase 3 rehabilitation was a priority in most settings, particularly in relation to planning physical activity in daily life. Psychosocial referrals were often made to patient associations or voluntary organisations to supplement the public healthcare services. While a few municipalities employed specific staff for such bridging activities, others placed little emphasis or priority on this aspect often providing support primarily to older adults.

A common feature in most settings was a final conversation, with a summary note sent to the patient’s GP at the end of the course including a treatment plan. In some settings, clinicians accompanied patients to their new activities or provided follow-up support after end of course.

### Phase 3 rehabilitation

The GPs were the primary clinicians and coordinators throughout the entire patient pathway, especially with responsibility for phase 3 rehabilitation. While the summary note sometimes prompted GPs to make proactive telephone calls to patients after discharge, it was often the patients themselves who had to make contact.

Phase 3 rehabilitation involved annual GP visits as follow-up on treatment, medication, the recovery process, and the maintenance of lifestyle changes. It also included managing referrals for further rehabilitation such as the regional hospital department providing lifestyle intervention programme, however this option was not mentioned by anyone. As psychologists are not part of the publicly funded health services, the GPs sometimes provide individual counselling as psychological support.

GPs perceived attendance and the quality of phase 2 rehabilitation as key factors influencing patients’ need for information and additional intervention. Patients were perceived to rely primarily on their self-management skills alongside support from community-based initiatives and activities. In this context, the Danish Heart Association was seen as an important collaborator and patients were encouraged to become members in order to gain access to information, opportunities for discussion and local activities.

Table 2 summarises the study’s key findings of the study, highlighting challenges and opportunities improving of CR pathways.

**Table 2:**
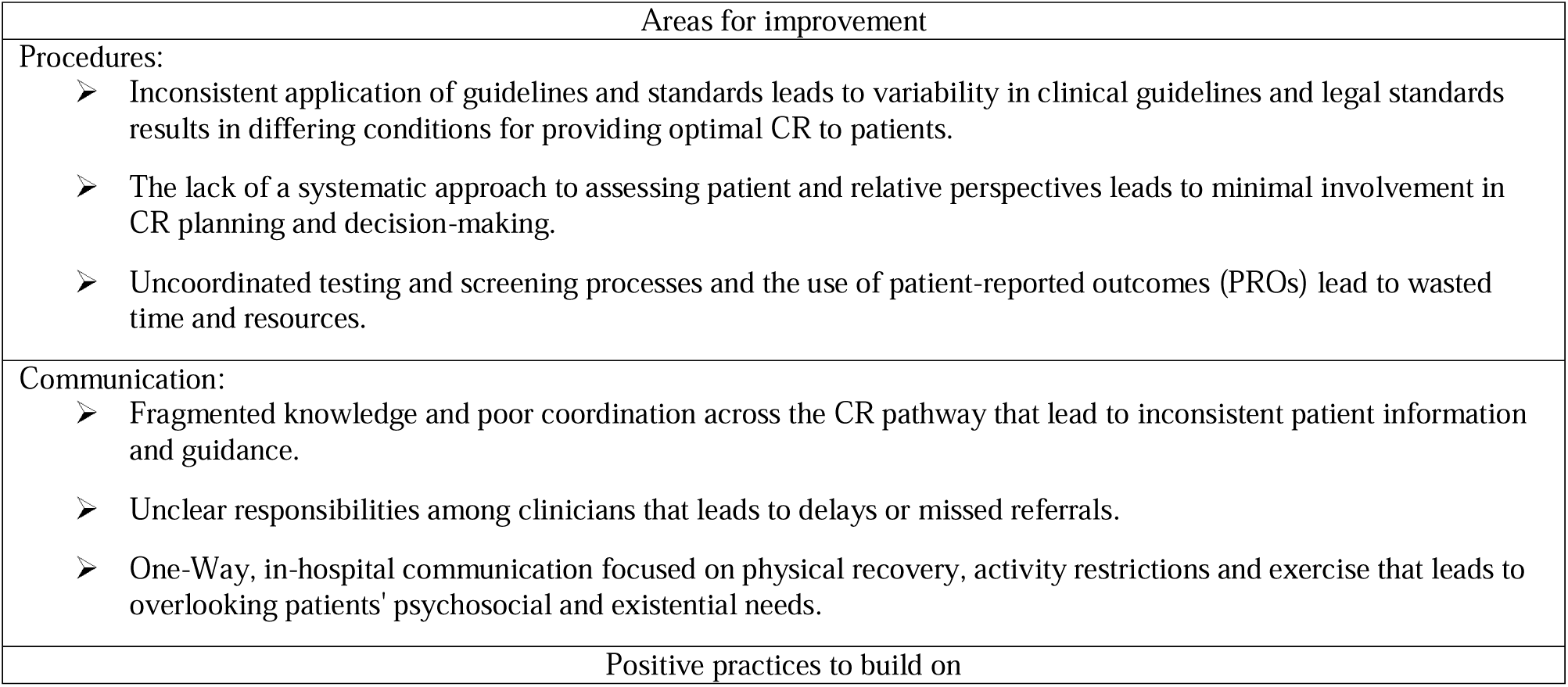
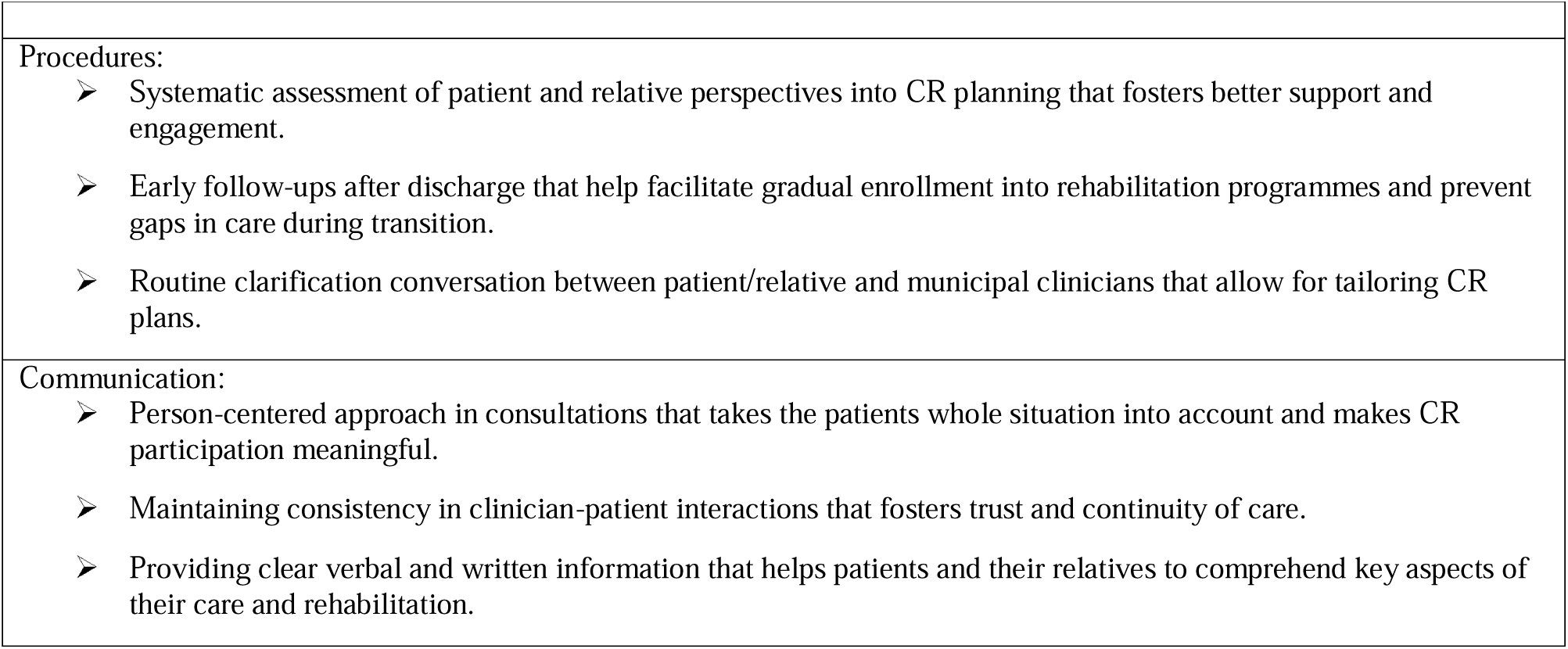
Summary of challenges and opportunities for CR innovations.

| Areas for improvement |  |
| --- | --- |
| Procedures: |  |
| ➤ | Inconsistent application of guidelines and standards leads to variability in clinical guidelines and legal standards results in differing conditions for providing optimal CR to patients. |
| ➤ | The lack of a systematic approach to assessing patient and relative perspectives leads to minimal involvement in CR planning and decision-making. |
| ➤ | Uncoordinated testing and screening processes and the use of patient-reported outcomes (PROs) lead to wasted time and resources. |
| Communication: |  |
| ➤ | Fragmented knowledge and poor coordination across the CR pathway that lead to inconsistent patient information and guidance. |
| ➤ | Unclear responsibilities among clinicians that leads to delays or missed referrals. |
| ➤ | One-Way, in-hospital communication focused on physical recovery, activity restrictions and exercise that leads to overlooking patients' psychosocial and existential needs. |
| Positive practices to build on |  |

|  |
| --- |
| Procedures: |
| ➤ Systematic assessment of patient and relative perspectives into CR planning that fosters better support and engagement. |
| ➤ Early follow-ups after discharge that help facilitate gradual enrollment into rehabilitation programmes and prevent gaps in care during transition. |
| ➤ Routine clarification conversation between patient/relative and municipal clinicians that allow for tailoring CR plans. |
| Communication: |
| ➤ Person-centered approach in consultations that takes the patients whole situation into account and makes CR participation meaningful. |
| ➤ Maintaining consistency in clinician-patient interactions that fosters trust and continuity of care. |
| ➤ Providing clear verbal and written information that helps patients and their relatives to comprehend key aspects of their care and rehabilitation. |

## Discussion

This study provides the first comprehensive overview of CR pathways following cardiac surgery in Denmark. Overall, our findings revealed minimal psychological support, patient involvement and collaboration with patients and their relatives in developing flexible, tailored CR plans. A key finding was a significant gap in care during the transition from hospital-based phase 1 to municipality-led phase 2 CR. This gap disrupted the continuity of the rehabilitation process, potentially causing significant setbacks in patients’ recovery. The considerable variation in referral processes and CR delivery observed in our study appears to stem in part from the lack of a clear legal framework and inconsistent guidelines.

Our findings emphasize the need for a more interactive, person-centred approach in CR, particularly during discharge consultations. Currently, these consultations primarily focus on providing information, without systematically addressing patients’ and relatives’ preferences, needs, or resources (33). While information is essential for preparing patients, a collaborative approach, such as shared decision-making, could improve CR engagement and outcomes of rehabilitation (34).

This study contributes to a better understanding of the CR pathway, enabling clinicians to provide consistent, accurate information and engage patients in discussions about CR options (35). The integration of patient perspectives could help overcome criticisms of previous research, which highlighted the lack of systematic methods to incorporate patients’ unique needs into treatment plans (33).

Furthermore, the systematic involvement of patients in designing the CR pathway could lead to more flexible and individually tailored approaches, particularly benefiting underserved groups and individuals requiring additional emotional and social support (14, 36–38). For successful implementation, clinicians must develop communication, listening, and negotiation skills, and tools and training can further enhance person-centred CR practices (39).

Transitions of care between settings are recognized as vulnerable periods for patients and their relatives (33). Our study identified a critical care gap during the transition, which was often prolonged, this may be due to poor coordination of handover and follow-up leaving patients with unresolved concerns and inadequate support (40). Dropout rates from CR are notably high during this period (5), highlighting the need for targeted interventions to improve patient retention.

A variety of transitional care programmes and services such as discharge planning, care coordination and case management have shown to be important to focus on patients with complex needs or vulnerabilities during the transfer from one type and setting of care to another (41). In Denmark, some initiatives have been developed to bridge the gap in transitional services from hospital to outpatient CR. Recently, a screening tool has been developed to identify patients at risk of dropping out of phase 2 CR, offering them individualised support, including a dedicated contact person and systematic follow-up and ensure enrolment (42). However, the intervention is not developed to work across sectors. Another Danish study provided a scheduled follow-up for patient after cardiac surgery in a medical student-led clinic, however this intervention was focused on physical complications and was time-limited with the primary goal of serving as a learning opportunity (43). In the UK a transitional intervention called the Cardiac Transitional Rehabilitation Using Self-Management Techniques (Cardiac TRUST) was developed and was promising in enhancing physical activity prior to outpatient CR (44). However, the intervention did not involve relatives nor integrate or adapt to patient perspectives. The need for such targeted transition strategies is becoming increasingly urgent because of factors such as shorter hospital stays, an ageing population with complex health needs, and the underrepresentation of certain patient groups in CR, including women, individuals with low socioeconomic status, and those facing logistical barriers, underscore the importance of structured and individualised support following discharge (45).

Clinicians in our study faced challenges due to the absence of updated national clinical guidelines on CR. This lack of regulatory consistency makes it difficult for clinicians to create rehabilitation programmes that are both standardised and adaptable to individual patient needs, despite European CR guidelines recommending a high level of patient involvement.

National legislation ensures universal rehabilitation coverage, essential for planning (37, 46), but aligning patient needs with these frameworks remains challenging (17). Future guidelines should incorporate patient perspectives to address complex healthcare needs, avoiding a narrow focus on technical aspects (47). Redesigning services with a focus on patient involvement in care planning will enhance rehabilitation outcomes (48). A key barrier is the tension between institutional policies and person-centred care. Balancing institutional requirements with individualized approaches will be vital for effective CR programmes (17).

The findings of this study highlight key areas for refining the current CR pathway and emphasize the need for a person-centred redesign, including improved sector transitions and revised guidelines.

### Strengths and limitations

A strength of this study is its use of multiple methods to provide both breadth and depth in describing touchpoints along the care pathway. As data were collected from specific healthcare settings in central and northern Denmark, they may not fully represent the diversity of CR pathways in other regions or countries. In addition, the cross-sectional nature of the study captures current practices but does not account for potential changes in rehabilitation pathways nor descriptions of the active ingredients of intervention to innovate changes in different service delivery contexts.

The metro-mapping methodology was originally developed for shared decision-making and planning in cancer care pathways (21). It has provided a useful methodological approach to unpack service delivery and experience level data in the context of CR pathways of care (56). This approach provides a valuable way to visualise the CR pathway, identifying points were patients and professionals interact to make decisions about care, and implement plans to monitor and enhance care processes (49). Metro mapping seems to be a useful method for designing components of a complex research interventions (50).

The use of multiple methods for triangulation, together with input from patient partners and participants from different healthcare sectors, strengthened the reliability of the findings by integrating both experiential knowledge and professional perspectives (51).

### Future perspectives

This study lays the groundwork for meaningful innovations in CR to enhance patient engagement, recovery, and continuity of care.

We have identified three key priorities: 1) improving communication of rehabilitation options and decision points to support shared decision-making, using tools such as adaptable patient decision aids; 2) bridging the existing gap in care through standardized transition protocols between hospital and outpatient settings, digital follow-up tools, and coordinated multidisciplinary care pathways; and 3) exploring flexible delivery formats—such as mHealth solutions, home-based CR, and hybrid models to enhance accessibility and engagement.

### Conclusion

This study provides a comprehensive overview of rehabilitation pathways and standard programmes following cardiac surgery in Denmark, highlighting significant variability and challenges. Our findings emphasise the need for a person-centred redesign of the cardiac rehabilitation pathway with improved sector transitions. Our findings underscore the importance of clear legal frameworks, consistent guidelines, and underscores the rationale for implementing individualised decisions about CR, given the complexity and diversity of healthcare services and needs, including those of the elderly, frail persons and those with co-morbidities. Strengthening these areas could promote greater patient engagement, reduce variability in service delivery, and ultimately lead to better patient outcomes and experiences.

## Supporting information

**S1 File: Standards of Reporting Qualitative Research (SRQR) checklist** (pdf)

**S2 File: Guidance for Reporting Involvement of Patients and the Public in Research-Short Form (GRIPP2-SF)**(pdf)

**S3 File: Matrix of involvement of patient partners in the research process** (pdf)

## Conflicts of interest

The authors declare no conflicts of interest.

## Funding

This work was support by the Novo Nordic Foundation (<u>Project Grants for Studies on IntegratedCare Pathways - Novo Nordisk Fonden</u>) Grant no. 0083480 received by ISM and BST. The funders had no role in study design, data collection and analysis, decision to publish, or preparation of the manuscript.

## Data Availability

All relevant data are within the manuscript and its figure.

## Acknowledgement

We thank the managers for granting access to healthcare staff, the participants for sharing their experiences, and the patient partners Vicky Elisabeth Vallund; Helle Kjær Sørensen; Mogens Møller Hansen; Jesper Gad Christensen and Preben Nielsen for their involvement in the research process.

## Author contributions

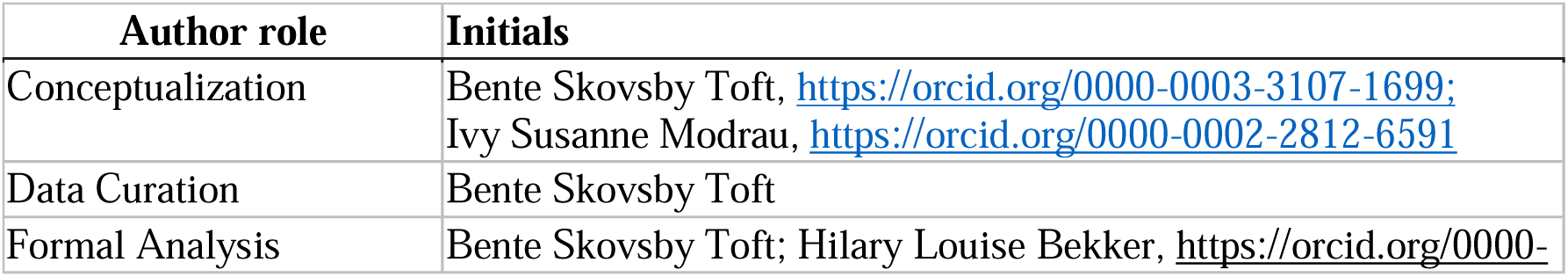
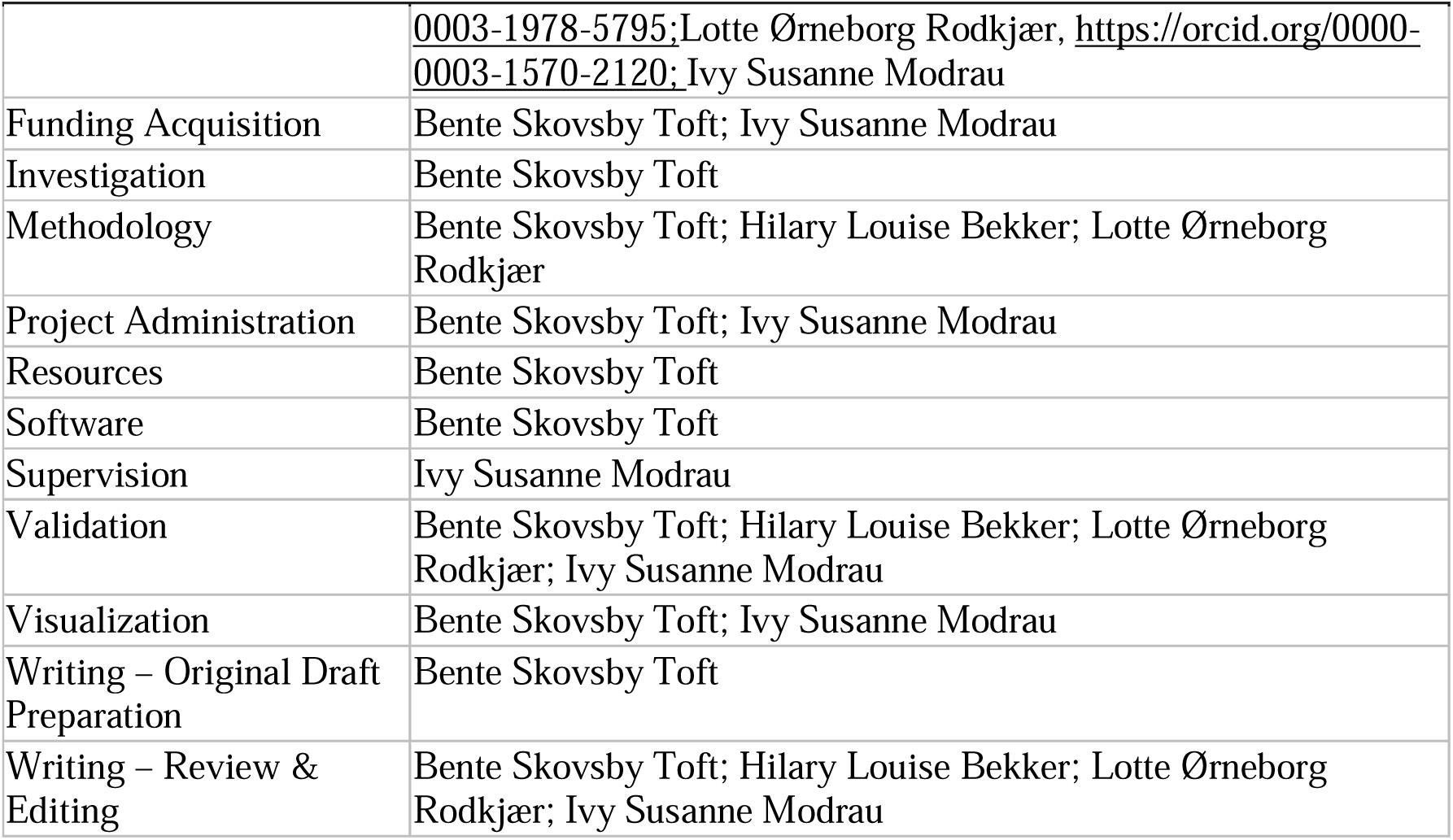

## Notes

### Competing Interest Statement

The authors have declared no competing interest.

### Funding Statement

Yes

### Author Declarations

The study was registered as an internal inventory of research projects with the Danish Data Protection Agency, Central Denmark Region, Regional Office (Skottenborg 26, DK-8800 Viborg), and the registration was confirmed (no. 1-16-02-304-24, 19 August 2024). The Committees on Health Research Ethics of the Central Denmark Region (Regional Secretariat Legal Office, Skottenborg 26, DK-8800 Viborg) were contacted and confirmed that, in accordance with the Consolidation Act on Research Ethics Review of Health Research Projects, Consolidation Act No. 1268 of 28 November 2024, Section 14(1), the study could be conducted without approval from the Committees. Ethical oversight was therefore waived by the ethics committee.

